# Strengthening Community Roots: Anchoring Newcomers in Wellness and Sustainability (SCORE!): A protocol for the co-design and evaluation of a healthy active living program among a newcomer community in Canada

**DOI:** 10.1101/2023.07.06.23292304

**Authors:** Gita Wahi, Sujane Kandasamy, Shrikant Bangdiwala, Andrea Baumann, Mary Crea- Arsenio, Dipika Desai, Kathy Georgiades, Fatimah Jackson-Best, Matthew Kwan, Patricia Montague, Bruce Newbold, Diana Sherifali, Amanda Sim, Russell J. de Souza, Sonia S. Anand, the SCORE! Research team

## Abstract

**BACKGROUND:** The burden of childhood obesity and cardiometabolic risk factors affecting newcomer Canadians living in lower socioeconomic circumstances is a concerning public health issue. This paper describes Strengthening Community Roots: Anchoring Newcomers in Wellness and Sustainability ([SCORE!](https://okanagan.mcmaster.ca/initiatives/score/#tab-content-ov)), an academic-community research partnership to co-design interventions that nurture and optimize healthy activity living (HAL) among a community of children and families new to Canada in Hamilton, Ontario, Canada.

**METHODS/DESIGN:** Our overarching program is informed by a socio-ecological model, and will co-create HAL interventions for children and families new to Canada rooted in outdoor, nature-based physical activity. We will proceed in three phases: Phase 1) synthesis of existing evidence regarding nature based HAL interventions among children and families; Phase 2) program development through four data collection activities including: i) community engagement activities to build trustful relationships and understand barriers and facilitators, including establishing a community advisory and action board, qualitative studies including a photovoice study, and co-design workshops to develop programs; ii) characterizing the demographics of the community through a household survey; iii) characterizing the built environment and HAL programs/services available in the community by developing an accessible real-time systems map; and iv) reviewing municipal policies relevant to HAL and sustainability; leading to Phase 3) implementation and evaluation of the feasibility of co-designed HAL programs.

**CONCLUSION:** The etiology of childhood obesity and related chronic diseases is complex and multifactorial, as are intervention strategies. The SCORE! program of research brings together partners including community members, service providers, academic researchers, and organizational leaders to build a multi-component intervention that promotes the health and wellness of newcomer children and families.

## INTRODUCTION

Obesity, diabetes and cardiovascular disease (CVD), known as non-communicable diseases (NCDs), collectively account for more than half of the global burden of disease among adults.^1^ Unfortunately with the growing burden of childhood obesity, cardio-metabolic complications are increasing among children and reducing life expectancy.^2^ In Canada, 32% of children aged 5-11 years are overweight or obese.^3^ In high-income countries, the burden of childhood obesity disproportionately affects those of lower socioeconomic status.^4^ Furthermore, racialized children in high income countries have more obesity and related cardiometabolic complications, including type 2 diabetes, compared to non-racialized children and youth.^5, 6^

The burden of childhood obesity and cardiometabolic risk factors affecting Canadians who have immigrated and are living in lower socioeconomic circumstances is an emerging issue.^7^ The growth in Canada’s population is led by immigration, where three-quarters of the 1% increase in population each year is accounted for by newcomers to Canada.^8^ From the 2021 Canadian census, 31.5% of children, had at least one parent born outside of Canada.^9^ Children from immigrant families are at higher risk for living in lower socioeconomic circumstances,^10^ which impacts their risk for obesity and its associated health consequences.^4, 11^ The associated challenges influenced by their socioeconomic position include language and employment barriers^12^, reduced recreational opportunities^13^, and housing opportunities^14^ which usually leads to fewer scheduled and unscheduled opportunities for physical activity^15^ and greater consumption of energy-dense, ultra-processed foods.^14, 16^

Healthy active living (HAL) refers to physical, mental and spiritual practices that promote health and wellbeing.^17^ Connections to nature through accessing parks, gardens, and playgrounds provide children and families with opportunities for HAL practices such as physical activity and social connectedness.^18^ Exposure to outdoor greenspace and nature-based activities, like community gardening^19^, is associated with higher levels of mental well-being and physical health.^20, 21^ Given the circumstances in which newcomer children in low-income settings may live, including reduced opportunities for physical activity due to urban living environments, we posit that early exposures and opportunities to outdoor greenspace and nature-based activities may improve newcomer children and families’ mental and physical health. Further, we aim to amplify the voices of those with lived experiences, and work in partnership with newcomer communities to co-create optimal HAL solutions.

This paper describes the academic-community research partnership: *Strengthening Community Roots: Anchoring Newcomers in Wellness and Sustainability* ([SCORE!](https://okanagan.mcmaster.ca/initiatives/score/#tab-content-ov)) to co-design interventions that seek to nurture and optimize healthy activity living to prevent obesity and related crdio- metabolic risk factors among a community of newcomer children and families in Hamilton, Ontario, Canada.

## METHODS/DESIGN

### Characteristics of population

The initial community of focus for SCORE! is the Riverdale neighbourhood of Hamilton, Ontario. The characteristics of the families in this community can be described as half of the residents (50%) identifying as a visible minority, intersecting with a quarter (25%) families immigrating to Canada in the last 20 years, a quarter (26%) identify as low income, as defined by Statistics Canada low-income measures.^22, 23^ The neighbourhood includes communities of South Asian, Southeast Asian, and Middle Eastern heritage with the most common linguistic preferences being Punjabi and Arabic.^23^

### Setting/Location

Community spaces in Riverdale include local public schools, playgrounds, and a city run recreation centre attached to the local elementary school. The primary residential dwellings are high-rise apartment buildings with limited access to safe greenspace for local families use. (Figure 1)

**Fig 1:**
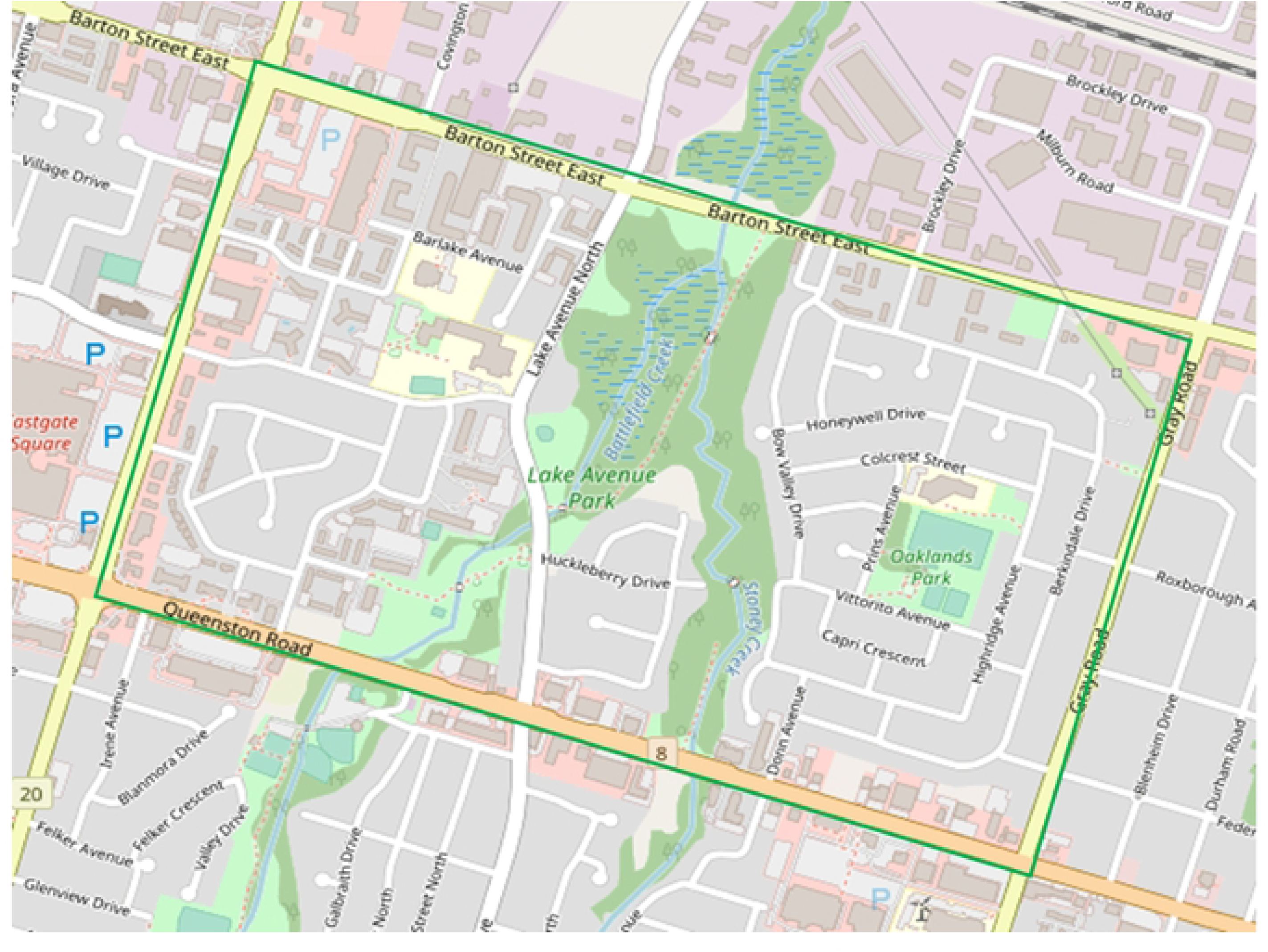
Map of Riverdale neighbourhood of Hamilton, ON, Canada.

### Theoretical framework and study objectives

The etiology of obesity is complex and multifactorial, as are prevention strategies.^24^ Previous approaches to prevent childhood obesity that exclusively took an individual-level approach and emphasized the sum of the individual balance of energy consumption (i.e., diet) and energy expenditure (i.e., physical activity), failed to acknowledge the intricate interplay of biology, psychology, socioeconomics and the built environment.^25^ A socio-ecological model allows for a broad approach to understanding the factors that contribute to childhood obesity and the interplay between various key components including the individual, relationships with family and peers, community factors and socio-cultural environments, built environment, and society.^26–28^

Using the socio-ecological framework to understand factors contributing to overweight and obesity, the overarching program goals of SCORE! are to co-create components of a HAL intervention for children and their families in three phases (Figure 2): Phase 1) synthesis of existing evidence regarding nature-based HAL interventions among immigrant children and families; Phase 2) program development through data collection activities at each level of the framework including: i.) community engagement activities to build trustful relationships and understand barriers/ facilitators to HAL including establishing a community advisory and action board, and qualitative studies including a photovoice study and co-design workshops; ii.) understanding the composition of the community and their access to health services and opinions about their community through a household survey; iii.) characterizing the built environment and HAL services available in the community by building a real-time, accessible systems map; and iv.) policy briefs will be prepared and shared with the key stakeholders involved in the policy-making processes with a policy round table; leading to Phase 3) the implementation and evaluation of co-designed HAL interventions to inform future SCORE! interventions.

**Fig 2:**
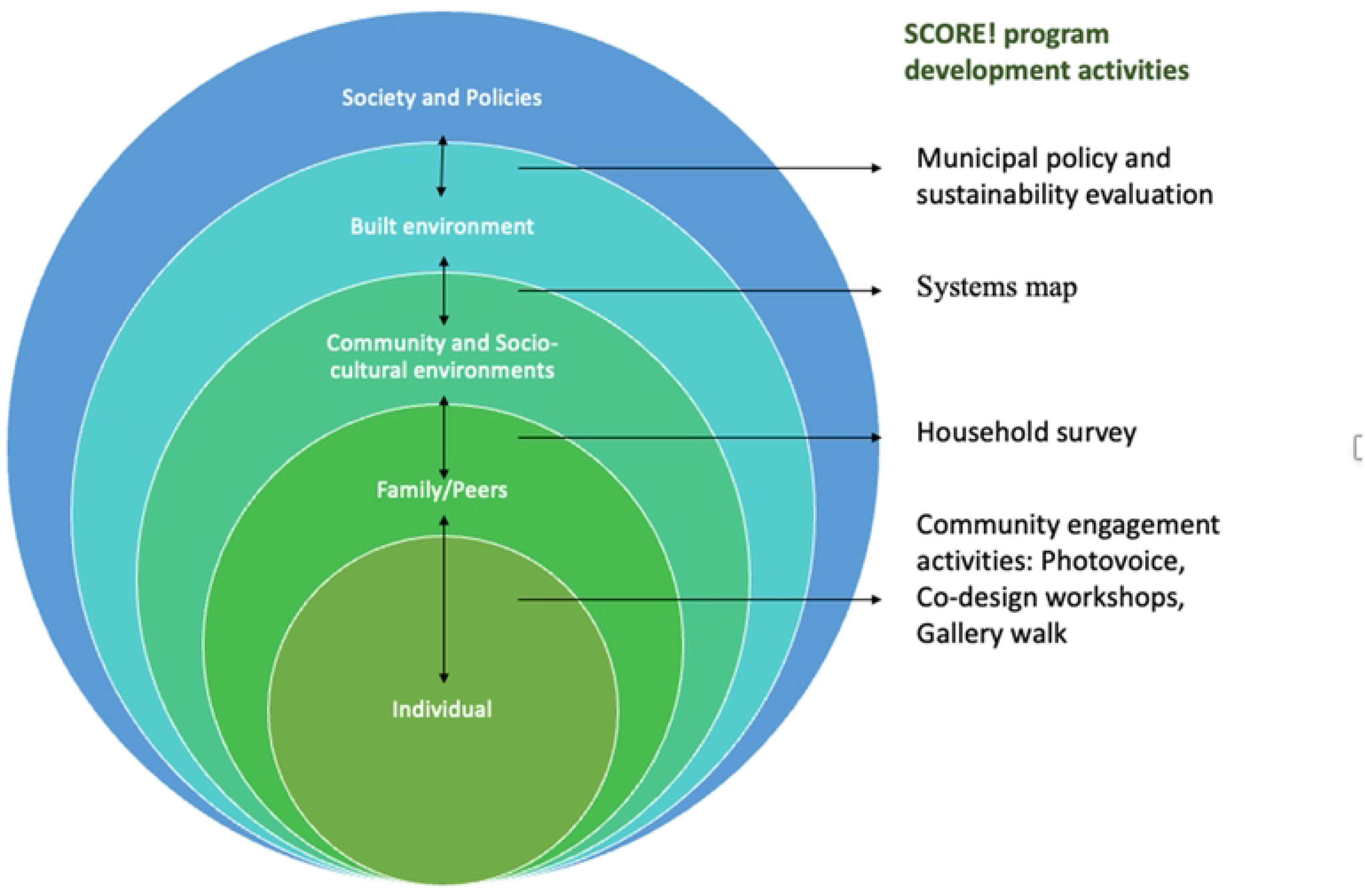
Socio-ecological framework adapted for SCORE! program development activities.

### Study activities

***Phase 1. Evidence Syntheses*** are being conducted to include scoping and systematic reviews of topics that encompass immigrant children and families, nature-based programs, outdoor healthy activity living interventions, and chronic disease prevention and management.

**Phase 2. Program Development**

a. *Community Engagement* Strong partnerships and meaningful engagement with the community residents, organizations, schools, and existing community groups within the community are crucial components for the design and evaluation of interventions to improve healthy active living among children and families. A community advisory and action board (CAB) has been established, with key members including local families, community leaders, and local service organizations to provide community oversight and input into all aspects of the project. Furthermore, understanding facilitators of and barriers to HAL programs from the perspectives of community members is central to formative work. Three activities will be inaugural efforts to better characterize these perspectives:
  <u>i) Photovoice qualitative study</u> aims to engage local families of diverse ancestry with a focus on the three largest groups: South Asian, Southeast Asian, Middle Eastern communities and organizations that work closely with children, youth, and families in the Riverdale area to develop an understanding of community perspectives and experiences of local greenspace and nature access. Photovoice is a qualitative method used in community-based participatory research to document and reflect personal experiences.^29^ The process is empowering, flexible, and embedded in grassroots social action to enhance community engagement.^30^
  <u>ii) Gallery walk activity</u> is an inquiry-based approach that allows participants to freely express their opinions and perspectives on a series of questions in a group or classroom format. Here children are engaged in school classrooms (kindergarten to grade 8) to describe physical and outdoor activities and programs they enjoy participating in, and to indicate which types of new activities they would like to explore and experience within their community, and the barriers and facilitators children identify as affecting their participation in activities and programs.
  <u>iii) Co-Design workshops</u> will engage a broad sample of representation to co-design HAL interventions using experience-based co-design (EBCD) because of its suitability for multiphase projects and in designing interventions.^31^ Key study partners will be community members, community partners including individuals who work with an organization in the community, and organizational leaders. To include representation from the community and service users, speaking English is not a requirement to participate in the workshops as interpreters are used as needed. The projected output will include an understanding of key barriers and facilitators to HAL and prototypes of interventions that will address the facilitators and barriers to HAL. The co-designed prototypes will be adapted to local context and resources by the research team and community partners and evaluated in Phase 3.
b. *<u>The SCORE! Household Survey</u>* will characterize the composition of the community through a cross-sectional survey. This information at the household level is not available from the Statistics Canada National Census, national surveys, or administrative health data. All households in the Riverdale geographic boundaries will be eligible and only one adult member of each household will complete the survey on behalf of the household. Key data collected will include household member composition including the number, sex/gender, and ages of adults and children living in the household along with factors from the household including health literacy, access to health services, and household perception of: 1) social cohesion, 2) social capital, 3) community pride, and 4) city-neighbourhood relationships. The sampling approach strives to ensure the same number of households within each census unit of the dissemination area (the Riverdale geographic boundary) to generate a sample representative of the community including that of immigration status, race/ethnicity, languages spoken, and income status.^32^ We anticipate that some residents of Riverdale will have lower English proficiency. To help increase recruitment, surveys are translated into multiple languages commonly spoken in the community including Arabic, Urdu, Farsi, and Punjabi. SCORE! multi-lingual team members will canvas door-to-door in the apartment buildings to speak directly with residents and translate/interpret recruitment materials.
c. *<u>Systems Map</u>* will characterize the built environment and HAL programs and services by documenting key formal and informal spaces in the city that offer programs and activities for children and families. To amplify the impact and utility of the systems map for the Riverdale residents, semi-structured interviews with community leaders will be conducted to assist in identifying spaces, services, resources, and programs within, or near, and available to the Riverdale community. These interviews will inform the development of the systems map by identifying and describing relevant services and resources, which will populate a map using an online GIS systeme. The SCORE! systems map will have the capability of being automatically updated as new source data are updated and changes. The systems map is being translated into the three most common languages in the Riverdale community.
d. *<u>Policy Round Table</u>:* Policy briefs will be prepared and shared with the key stakeholders involved in the policy-making processes that influence the primary community of interests opportunities for HAL. This will include Hamilton Public Health, City of Hamilton, Hamilton Wentworth District School Board, and Hamilton Catholic School Board, advocacy organizations, and other key actors. Policy round table discussions will be the starting point and lead to development of the framework for the assessment of HAL-related policy and implementation, as well as the extent to which policy outcomes align with the priorities of different stakeholder groups.

***Phase 3.* Implementation and evaluation of the feasibility of co-designed HAL interventions** will be guided by the RE-AIM framework for public health intervention evaluation.^33^ This framework includes measurement of the reach, effectiveness, adoption, implementation, and maintenance of the interventions.^33^ This will be a pre-post study design to pilot and evaluate activities that have been co-designed within the SCORE! program platform. Key feasibility objectives for this piloting exercise will include: 1) Recruitment (e.g., how many families expressed interest and consent) to the activity; 2) Uptake of the intervention (e.g., how many people who consent attend the activities); 3) Among those who attend the events, the percent that complete baseline and follow-up questionnaires to measure nutrition, physical activity, physical literacy, and sleep (of both child and parents) at baseline and follow-up questionnaires. 4) Evaluate the acceptability, barriers and facilitators to participating in the HAL interventions.

### Knowledge Exchange

Knowledge exchange will occur using a multi-pronged and integrated knowledge translation strategy that builds on existing community partnerships and integrates new networks that we will cultivated through this project knowledge exchange will occur. We first plan to co-synthesize key learnings with the leadership of the advisory councils and committees. This will be presented to all partner organizations via regular knowledge exchange meetings. During these sessions, we will review progress, corroborate learnings, explore divergences, and discuss the next steps. At the end of the project activities, we will bring all parties together for a meeting where we will review key lessons learned and develop a road map for implementation. Second, existing communication channels and social media avenues will be leveraged to disseminate key messages, learnings, and results throughout the project. The McMaster Okanagan Committee, which is charged with sustainability initiatives for the university campus and the surrounding city will manage our social media posting and website (https://okanagan.mcmaster.ca). This will harness and amplify the voices of the participants (i.e., priority populations) by having those interested lead the e-journal pieces, select meeting topics, and represent their communities of interest on our governance committees. These activities will encourage active involvement of residents through sharing their stories, directing their narratives, and building capacity for leadership. Figure 3 illustrates SCORE!’s governance structure.

**Fig 3:**
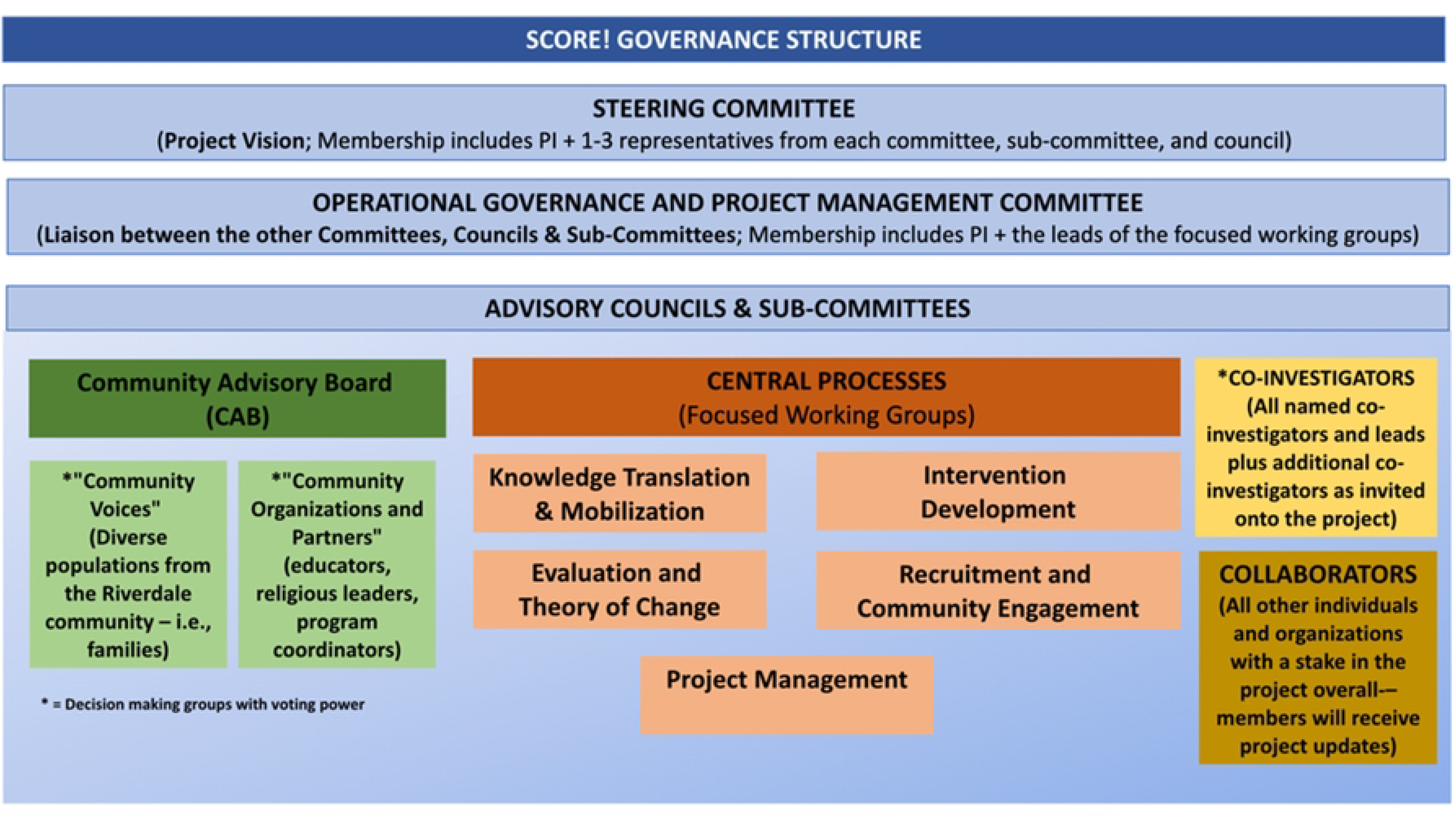
Governance model of SCORE!

### Instruments and Data tools

We will use validated data collection instruments and questionnaires in the Household survey (including questions on health literacy, access to health services, and household perception of social cohesion, social capital, pride living in their community, and city- neighbourhood relationships) (Table 1a) and for evaluating the interventions (nutrition, physical activity, physical literacy, mental health and well-being, and sleep). (Table 1b)

**Table 1.**
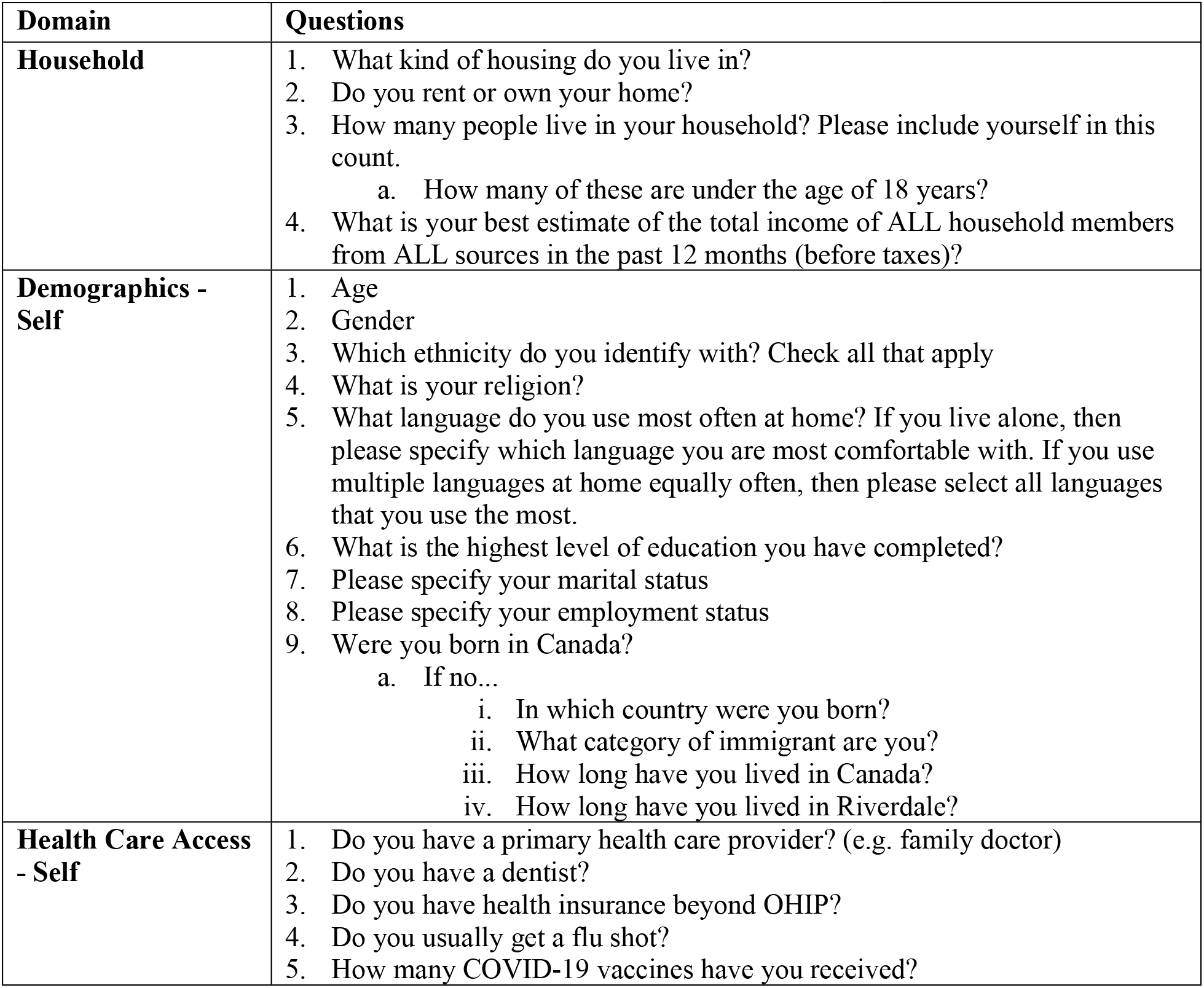
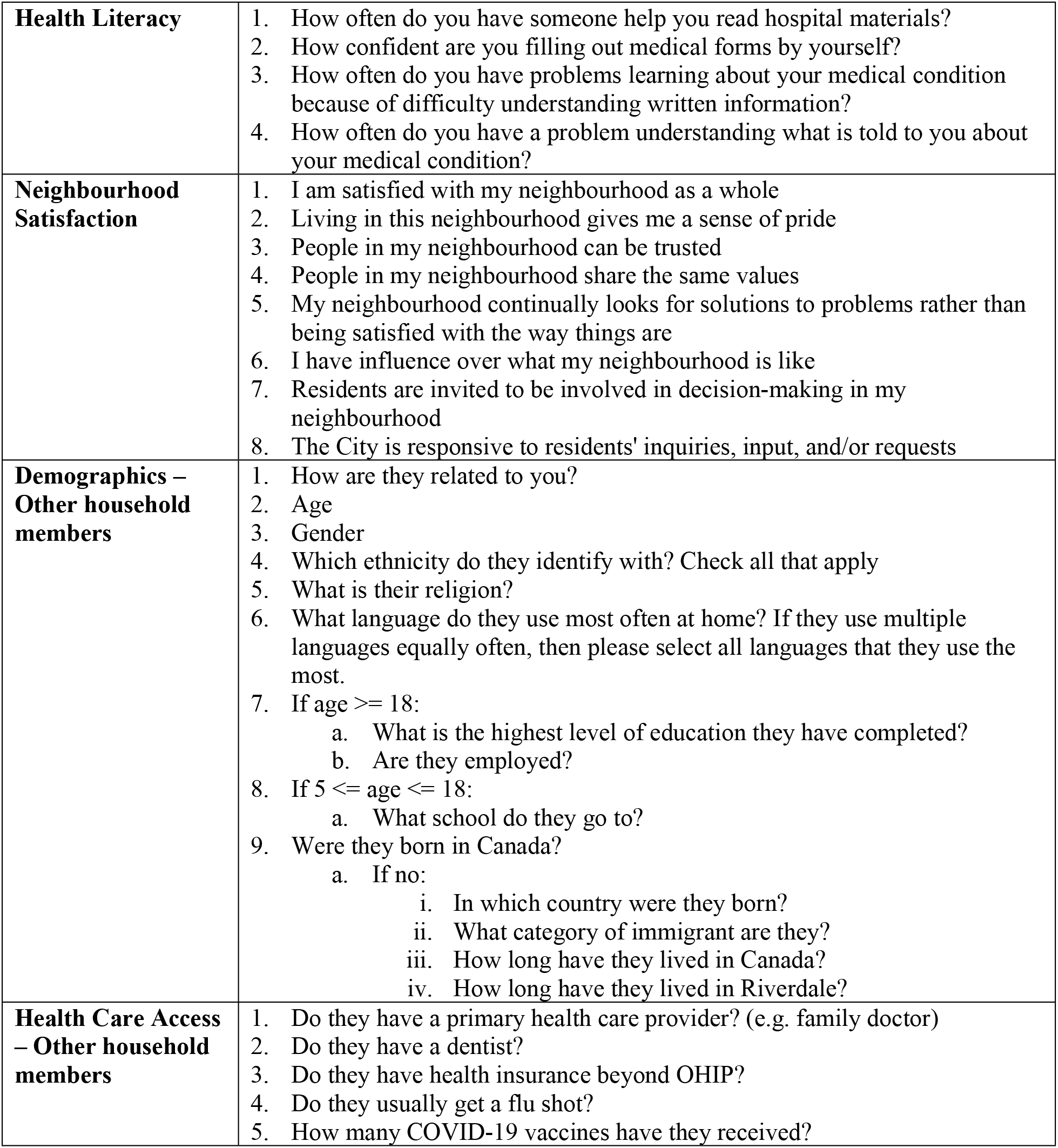
Data instruments for the household survey.

**Table 1b:** Data instruments for the pilot intervention implementation and evaluation.

| Domain | Questions |
| --- | --- |
| Household | 1. What kind of housing do you live in?<br>2. Do you rent or own your home?<br>3. How many people live in your household?<br>a. How many of these are under the age of 18 years?<br>4. What is your best estimate of the total income of ALL household members from ALL sources in the past 12 months (before taxes)? |
| Demographics | 5. How old are you?<br>6. What is your gender?<br>7. Which ethnicity do you identify with? Check all that apply<br>8. What is your religion?<br>9. What language do you use most often at home?<br>10. Adults only:<br>a. What is the highest level of education you have completed?<br>b. Please specify your marital status<br>c. Please specify your employment status<br>11. Children only:<br>a. What grade are you in at school? In which country were you born?<br>12. How many years have you lived in Canada? |
| Healthy Active Living - Parent | 1. How many people smoke in your household?<br>2. How often does your family eat out at a restaurant or order in food?<br>3. Over the past 7 days, on how many days were you physically active for a total of at least 60 minutes per day?<br>4. Over the last 7 days, on how many days was your child physically active for a total of at least 60 minutes per day? |
| Sleep | Please answer the following questions about your child's sleeping patterns over the past month:<br>1. What time do they usually get up in the morning?<br>2. What time do they usually go to bed?<br>3. How many hours of sleep do they usually get? |
| Physical Activity | 1. How long does your child do the following, on average? [answer separately for weekdays and weekends]<br>a. Total Screen time<br>b. Play Outside<br>c. Organized Sports |
| Diet | How often does your child eat each of the following in a typical week?<br>1. Deep fried food/snacks/fast food<br>2. Salty foods/snacks<br>3. Chocolate<br>4. Desserts/sweet snacks<br>5. Pop/soda<br>6. Nuts/seeds<br>7. Fruits<br>8. Fruit juices<br>9. Leafy green vegetables |

### Data management

Participant data will be anonymized and stored in a secure electronic data platform.

### Timeline

Study activities began in April 2022 and will continue through March 2024.

### Key Partnerships

Non-profits organizations (e.g., Green Venture), academic (McMaster University, Mohawk College), the City of Hamilton including recreational centres and public health, along with local schools including the school administration. Table 2 illustrates key partners by sector.

**Table 2:**
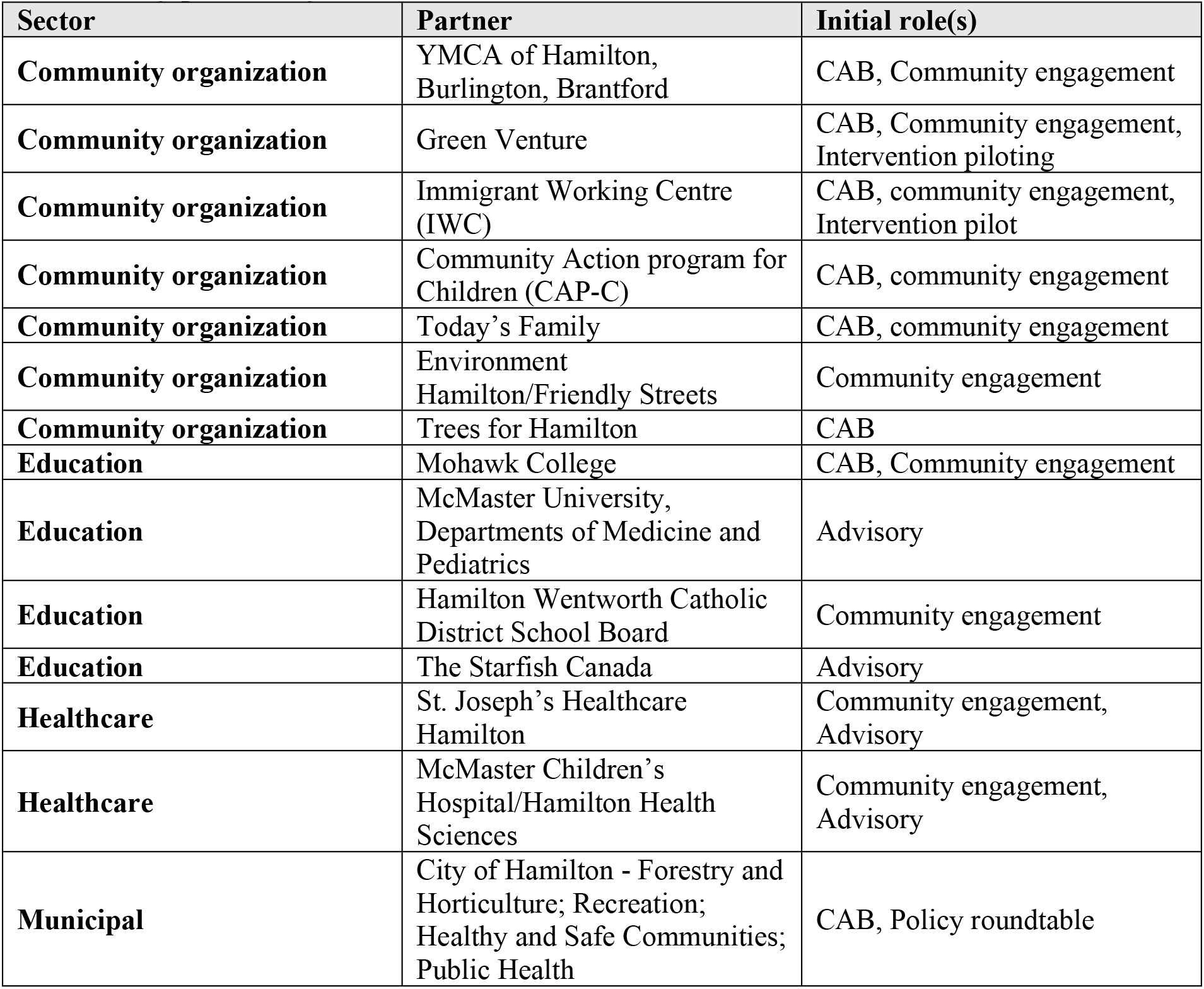
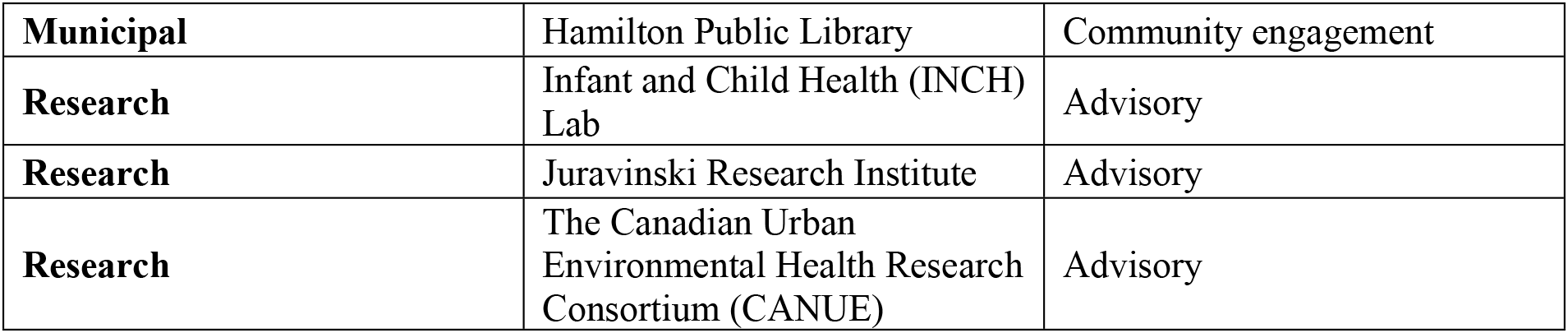
Key partners by sector

### Ethics

As the project involved extensive community engagement activities and co-design with community partners, indepth discussions were undertaken within the study team, with our community and funding partners, and our academic research ethics board with respect to in which settings and for which activities, a full description of research intent and informed consent was required. The activities undertaken as part of SCORE! were guided by the Tri Council Policy Statement-2: Ethical Conduct for Research Involving Humans.^34^ For each research study activity described above separate protocols have been approved if applicable by the Hamilton Integrated Research Ethics Board. When personal information were collected community participants were asked to provide informed consent.

## DISCUSSION

This protocol outlines the inaugural phases of the development of the SCORE! program of research which will co-design nature-based, outdoor physical activity HAL interventions with a community of newcomer children and families in Ontario, Canada.

Obesity is a significant public health concern with complex and multi-factorial levers and barriers. The trajectory of growth in childhood is a symptom of a larger imbalance not merely at the level of the individual or even the family, but the built environment and economy which requires population-level approaches.^28^ The use of methods to co-create multi-level interventions has been shown to improve adherence and fidelity to programming. Thoughtful, inclusive, and sustainable engagement with multiple stakeholders is required for wholesome planning and intervention design. We estimate that this approach will take at least 2 years in the design phase.

There are unique components to this research protocol. The focus on the primary prevention of obesity and related health complications of a high-risk population, newcomer children and families^11^ comes at a time when the population growth in Canada is driven by global migration patterns. Understanding the unique needs and challenges of this community is important for ensuring adequate service provision no, and in the future. Further, the connection to nature and outdoor time highlights the rising concerns about a climate crisis that has direct connections to health. A systematic review by An *et al* describes a conceptual model that connects global warming to the obesity epidemic through the impact of fossil fuels, population growth, and industrialization as factors leading to changes in land use, agricultural productivity, and transportation. They demonstrate how these upstream factors should be factored into contributing to transitions in nutrition and physical activity to impact the obesity epidemic.^35^ Understanding local context and global levers will be important to drive meaningful health interventions.

Initial activities to launch SCORE!’s multi-level projects have tested operational processes leading to challenges, as well as highlighting the importance of community partnerships. Early challenges include navigating and adapting academic institutional processes for community-engaged research. Other challenging processes include navigating the complex web of community programs and services, which has demonstrated that public services are often siloed and not designed optimally to service the most vulnerable. The voices of the community members are infused through every aspect of this project. Community members in the CAB and other community engagement activities have been instrumental in project decision making including as examples the research agenda, choosing the most common non-English languages for translation of materials, where and how to hold events, and study recruitment.

While some challenges have presented themselves in the co-design of this intervention, we have developed collaborative relationship with key partners including the local schools, municipal government, non-profit agencies, and our academic research ethics boards.Furthermore, the creation of a Policy Round Table provides a vehicle to have multisectoral communication to enable our collective effort to establish the successful implementation and sustainability of the intervention we are designing.

This proposed work will be foundational to a platform of research that prioritizes a community-academic partnership to build public health interventions that aim to prevent obesity and promote nature-based physical activity, among a community of newcomer Canadians.

## Data Availability

No datasets were generated or analysed during the current study. All relevant data from this study will be made available upon study completion.

## SCORE! Research Team

**Investigators:** Sonia S. Anand, Shrikant Bangdiwala, Andrea Baumann, Jeffrey Brook, Mary Crea-Arsenio, Russell de Souza, Dipika Desai, Deborah Diliberto, Kathy Georgiades, Fatimah Jackson-Best, Sujane Kandasamy, Matthew Kwan, Bruce Newbold, Diana Sherifali, Amanda Sim, Gita Wahi. **Operations team:** Patty Montague (Program Manager), Natalie Campbell, Madison Fach, Abdul Naebi.

## Acknowledgements

Dr. Anand is supported by a Tier 1 Canada Research Chair in Ethnicity and CVD and Heart, Stroke Foundation Chair in Population Health, grant from the Canadian Partnership Against Cancer, Heart and Stroke Foundation of Canada and Canadian Institutes of Health Research

## Funders and Community Partners

Public Health Agency of Canada (PHAC); The Canadian Urban Environmental Health Research Consortium (CANUE); City of Hamilton, Community Action Program for Children (CAPC) (c/o Ghanwa Afach); Environment Hamilton/Friendly Streets; Faculty of Health Sciences (McMaster University); Global Health Program (McMaster University); Green Venture (Guiliana, Casimirri, Sheila Gutierrez, Heather Govender); Hamilton Public Library; Hamilton Wentworth District School Board (c/o Jeff Zwolak, Lake Avenue Elementary School); Hamilton Wentworth Catholic District School Board (c/o Morris Hucal); Immigrant Working Centre (IWC) (c/o Claudio Ruiz, Wasan Mohamad, Rosemary Aswani, Ahlam Mohammed); Infant and Child Health (INCH Lab at Brock University); Juravinski Research Institute; Master of Public Health Program (McMaster University); McMaster Children’s Hospital; McMaster Evidence Review and Synthesis Team (MERST); McMaster Okanagan Committee; McMaster University Department of Pediatrics, Mohawk College (Wendy Lawson), Novartis, The Research Institute of St. Joe’s Hamilton (Lehana Thebane); The Starfish Canada; Today’s Family (c/o Brenda Ferguson, Blaze Forgie); Trees for Hamilton (Myles Sergeant); YMCA of Hamilton, Burlington & Brantford (c/o Nicki Glowacki, Lily Lumsden, Parsa Memon).

## Administration

Loshana Sockalingam, Kathy Stewart.

## Students/trainees

Nora Abdalla, Sandi Azab, Shania Bhopa, Ashfia Chowdhury, Fatima Dawood, Rabbi Fazle, Gurlean Gill, Junaid Habibi, Salima Hemani, Margaret Lo, Baanu Manoharan, Saathana Mathirajan, Pranshu Muppidi, Abeerah Murtaza, Adonis Ng, Natasha Ross, Sohnia Sansanwal, Jaanuni Shanjith, Afraah Shirin, Tyler Soberano, Divya Tamilselvan, Sanya Vij, Aamina Zahid.

## Volunteers

Adan Amer, Albi Angjeli, Krishna Basani, Dania Buttu, Naisha Dharia, Paranshi Gupta, Araash Halani, Sarah Hassan, Senaya Karunarathne, Menhaz Munir, Lennisha Nagalingam, Melissa Pereira, Felicia Liu, Shanelle Racine, Dania Rana, Sachin Sergeant, Rosain Stennett.

## REFERENCES

1. Yusuf S, Reddy S, Ôunpuu S, Anand S. Global burden of cardiovascular diseases: part I: general considerations, the epidemiologic transition, risk factors, and impact of urbanization. Circulation. 2001;104(22):2746–2753.

2. Di Cesare M, Sorić M, Bovet P, et al. The epidemiological burden of obesity in childhood: a worldwide epidemic requiring urgent action. BMC Med. 2019/11/25 2019;17(1):212. doi:10.1186/s12916-019-1449-8

3. Roberts KC, Shields M, de Groh M, Aziz A, Gilbert JA. Overweight and obesity in children and adolescents: results from the 2009 to 2011 Canadian Health Measures Survey. Health reports. Sep 2012;23(3):37–41.

4. Blasingame M, Samuels LR, Heerman WJ. The Combined Effects of Social Determinants of Health on Childhood Overweight and Obesity. Childhood Obesity. 2023;doi:10.1089/chi.2022.0222

5. Amed S, Dean HJ, Panagiotopoulos C, et al. Type 2 diabetes, medication-induced diabetes, and monogenic diabetes in Canadian children. A prospective national surveillance study. Diabetes Care. 2010;33(4):786–791.

6. Hudda MT, Nightingale CM, Donin AS, et al. Patterns of childhood body mass index (BMI), overweight and obesity in South Asian and black participants in the English National child measurement programme: effect of applying BMI adjustments standardising for ethnic differences in BMI-body fatness associations. International Journal of Obesity. 2018;42(4):662–670.

7. Lane G, Farag M, White J, Nisbet C, Vatanparast H. Chronic health disparities among refugee and immigrant children in Canada. Applied Physiology, Nutrition, and Metabolism. 2018;43(10):1043–1058.

8. Government of Canada. Annual Demographic Estimates: Canada, Provinces and Territories, Analysis: Total Population. Accessed August 13, 2022. https://www150.statcan.gc.ca/n1/pub/91-215-x/2021001/sec1-eng.htm

9. Statistics Canada. Immigrants make up the largest share of the population in over 150 years and continue to shape who we are as Canadians. Government of Canada. Updated October 26, 2022. Accessed April 1, 2023. https://www150.statcan.gc.ca/n1/daily-quotidien/221026/dq221026a-eng.htm

10. Beiser M, Hou F, Hyman I, Tousignant M. Poverty, family process, and the mental health of immigrant children in Canada. Am J Public Health. 2002;92(2):220–227.

11. Wahi G, Boyle MH, Morrison KM, Georgiades K. Body mass index among immigrant and non-immigrant youth: Evidence from the Canadian Community Health Survey. Can J Public Health. 2014;105(4):e239–e244.

12. Alidu L, Grunfeld EA. A systematic review of acculturation, obesity and health behaviours among migrants to high-income countries. Psychology & Health. 2018/06/03 2018;33(6):724-745. doi:10.1080/08870446.2017.1398327

13. Lane G, Nisbet C, Johnson S, Candow D, Chilibeck PD, Vatanparast H. Barriers and facilitators to meeting recommended physical activity levels among new immigrant and refugee children in Saskatchewan, Canada. Applied Physiology, Nutrition, and Metabolism. 2021;46(7):797–807.

14. Ghosh-Dastidar B, Cohen D, Hunter G, et al. Distance to Store, Food Prices, and Obesity in Urban Food Deserts. American Journal of Preventive Medicine. 2014/11/01/ 2014;47(5):587-595. doi:https://doi.org/10.1016/j.amepre.2014.07.005

15. Lacoste Y, Dancause KN, Gosselin-Gagne J, Gadais T. Physical Activity Among Immigrant Children: A Systematic Review. Journal of Physical Activity and Health. 01 Oct. 2020 2020;17(10):1047-1058. doi:10.1123/jpah.2019-0272

16. Henderson A, Epp-Koop S, Slater J. Exploring food and healthy eating with newcomers in Winnipeg’s North End. *International Journal of Migration*, Health and Social Care. 2017;13(1):1–14. doi:10.1108/IJMHSC-06-2015-0022

17. Health Canada. Healthy Living. Government of Canada. Accessed April 1, 2023. https://www.canada.ca/en/health-canada/services/healthy-living.html

18. Flett MR, Moore RW, Pfeiffer KA, Belonga J, Navarre J. Connecting Children and Family with Nature-Based Physical Activity. American Journal of Health Education. Sep/Oct Sep/Oct 2010 2023–02-21 2010;41(5):292-300.

19. Al-Delaimy WK, Webb M. Community Gardens as Environmental Health Interventions: Benefits Versus Potential Risks. Current Environmental Health Reports. 2017/06/01 2017;4(2):252-265. doi:10.1007/s40572-017-0133-4

20. Tillmann S, Tobin D, Avison W, Gilliland J. Mental health benefits of interactions with nature in children and teenagers: A systematic review. J Epidemiol Community Health. 2018;72(10):958–966.

21. Van den Berg M, Wendel-Vos W, van Poppel M, Kemper H, van Mechelen W, Maas J. Health benefits of green spaces in the living environment: A systematic review of epidemiological studies. Urban forestry & urban greening. 2015;14(4):806–816.

22. Census Mapper. Canada Census 2021, Low Income Explorer (People in LIM-AT). Accessed May 12, 2023. https://censusmapper.ca/maps/3350#16/43.2321/-79.7574

23. Statistics Canada. Census Profile, 2021 Census of Population. Government of Canada. Accessed May 12, 2023. https://www12.statcan.gc.ca/census-recensement/2021/dp-pd/prof/details/page.cfm?Lang=E&SearchText=L8E%201J7&DGUIDlist=2021S05075370072.03&GENDERlist=1,2,3&STATISTIClist=1,4&HEADERlist=0

24. World Health Organization WHO. Obesity. Accessed April 19, 2021. https://www.who.int/health-topics/obesity#tab=tab_1

25. Finegood DT. The importance of systems thinking to address obesity. Obesity Treatment and Prevention: New Directions. Karger Publishers; 2012:123–137.

26. Richard L, Gauvin L, Raine K. Ecological models revisited: their uses and evolution in health promotion over two decades. Annu Rev Public Health. 2011;32:307–326.

27. Golden SD, Earp JAL. Social ecological approaches to individuals and their contexts: twenty years of health education & behavior health promotion interventions. Health Educ Behav. 2012;39(3):364–372.

28. Willows ND, Hanley AJ, Delormier T. A socioecological framework to understand weight-related issues in Aboriginal children in Canada. Appl Physiol Nutr Metab. 2012;37:1–13.

29. Wang CC. Photovoice: A participatory action research strategy applied to women’s health. Journal of women’s health. 1999;8(2):185–192.

30. Wang C, Burris MA. Empowerment through photo novella: Portraits of participation. Health education quarterly. 1994;21(2):171–186.

31. Fylan B, Tomlinson J, Raynor DK, Silcock J. Using experience-based co-design with patients, carers and healthcare professionals to develop theory-based interventions for safer medicines use. Research in Social and Administrative Pharmacy. 2021;17(12):2127–2135.

32. Sim A, Georgiades K. Neighbourhood and family correlates of immigrant children’s mental health: a population-based cross-sectional study in Canada. BMC Psychiatry. 2022/07/05 2022;22(1):447. doi:10.1186/s12888-022-04096-7

33. Glasgow RE, Harden SM, Gaglio B, et al. RE-AIM Planning and Evaluation Framework: Adapting to New Science and Practice With a 20-Year Review. Mini Review. Frontiers in Public Health. 2019-March-29 2019;7doi:10.3389/fpubh.2019.00064

34. Canadian Institutes of Health Research NSaERCoC, and Social Sciences and Humanities Research Council of Canada,. TriCouncil Policy Statement: Ethical Conduct for Research Involving Humans. Government of Canada; 2022. https://ethics.gc.ca/eng/policy-politique_tcps2-eptc2_2022.html

35. An R, Ji M, Zhang S. Global warming and obesity: a systematic review. Obesity Reviews. 2018;19(2):150–163. doi:https://doi.org/10.1111/obr.12624

